# Genome sequencing boosts diagnostic yield for the developmental and epileptic encephalopathies

**DOI:** 10.64898/2026.04.24.26351703

**Authors:** Jacob E. Munro, Harshini Thiyagarajah, Mark F. Bennett, Annie T.G. Chiu, Amy L. Schneider, Caitlin A. Bennett, Nico Lieffering, Talia Allan, Tom Witkowski, Rebekah V. Harris, Joshua Reid, Neblina Sikta, Sian Macdonald, Lucy Coulter, Yew Li Dang, Jennifer Kerkhof, Bekim Sadikovic, Piero Perucca, Samuel F. Berkovic, Soham Sengupta, Christy W. LaFlamme, Heather C. Mefford, Melanie Bahlo, Ingrid E. Scheffer, Michael S. Hildebrand

## Abstract

**Purpose:** Although most developmental and epileptic encephalopathies (DEEs) have a monogenic aetiology, routine clinical genetic testing is negative for 50% of patients. We hypothesized that the diagnostic yield could be increased in a large cohort of individuals with unsolved DEEs by applying genome sequencing along with enhanced variant analyses outside of coding regions.

**Methods:** We performed genome sequencing for 242 participants with DEEs negative on prior genetic testing. We interrogated single nucleotide variants (SNVs), indels, and structural variants in both established and candidate DEE genes. All variants of interest were reviewed, classified, and validated by a multidisciplinary team.

**Results:** A molecular diagnosis was discovered for 36/242 (15%) participants. The pathogenic or likely pathogenic variants comprised 26 SNVs and indels within coding regions, 9 structural variants, and 5 SNVs and indels in introns or non-coding genes. Variants of uncertain significance were detected in a further 10/242 (4%) participants.

**Conclusion:** Genetic diagnostic yield for individuals with unsolved DEEs improves with genome sequencing analysis. This increase reflects both the identification of structural and non-coding variants not detectable on exome or gene panel analysis, and the detection of variants in genes newly associated with DEEs.

## INTRODUCTION

Developmental and epileptic encephalopathies (DEEs) are the most severe group of epilepsies^1^. They are characterized by frequent seizures and/or epileptiform activity on electroencephalogram (EEG) associated with developmental plateauing or regression^2^. Onset is usually in infancy or childhood with drug-resistant seizures. DEEs result in lifelong disability, with immense social, psychological and financial impacts for individuals and their families. Patients typically have multiple morbidities including intellectual disability (ID), autism spectrum disorder and other psychiatric disorders, movement disorders, gastrointestinal problems and sleep issues. They also have a high mortality rate due to sudden unexpected death in epilepsy and status epilepticus^3^.

DEEs include over 15 defined electro-clinical syndromes, including Dravet syndrome, Lennox-Gastaut syndrome (LGS), epilepsy with myoclonic–atonic seizures (EMAtS), DEE with spike-wave activation in sleep (DEE-SWAS), and infantile epileptic spasms syndrome (IESS)^2^, although around one-third of patients with DEEs cannot be diagnosed with a specific DEE syndrome^4^. Syndromes are diagnosed by age of onset, specific seizure types, characteristic EEG patterns, comorbidities, clinical trajectories, and often specific genetic determinants. Over 90% of patients with Dravet syndrome have pathogenic variants in *SCN1A,* the gene encoding the alpha-1 subunit of the sodium channel^5^, while most other syndromes are genetically heterogeneous. While each individual DEE syndrome is rare, together they represent a significant disease burden, with an estimated cumulative incidence of 1 in 590 children^4^.

Although acquired causes such as hypoxic–ischemic encephalopathy or infection account for some cases, the majority of DEEs have a genetic basis^2^. Over 970 genes have been implicated as monogenic causes of DEE^6^ (see curated url: github.com/bahlolab/genes 4epilepsy), representing almost 5% of all protein-coding genes. These genes reflect the diverse molecular mechanisms underlying DEEs, including disruptions of ion channel function, synaptic transmission, neurotransmitter signalling, metabolic pathways, cellular growth, gene regulation, RNA splicing, and protein degradation^7^. The high frequency of *de novo* pathogenic variants causing DEEs is consistent with a predominantly monogenic architecture^8^, however, recent evidence shows that polygenic risk contributes to these disorders, influencing penetrance and severity^9,10^. Additional ultrarare variants also contribute to DEE phenotypes^11^. Nonetheless, the majority of undiagnosed DEEs are likely driven by pathogenic variants in a single gene of major effect.

A genetic diagnosis for individuals with a DEE is life-changing. It often ends a long diagnostic odyssey, alleviates parental guilt, and provides access to disease-specific resources, advocacy networks, and peer support^12^. Critically, establishing the molecular basis of disease informs prognosis, recurrence risk counselling, and management by guiding treatment and enabling enrolment in precision therapy trials. However, despite the extensive set of known DEE-associated genes, approximately half of individuals with DEEs remain without an etiological diagnosis following exome sequencing (ES) or multigene panels (MGP)^2^.

The current diagnostic gap likely reflects both undiscovered DEE-causing genes and the technical limitations of current approaches in detecting certain classes of genomic variation. Genome sequencing (GS) overcomes many of these technical limitations, as it captures a broader spectrum of variation that is missed by ES and panel methods, including structural variants, repeat expansions, and non-coding variants. Previous work on a small cohort of 44 patients with DEE achieved an additional diagnostic yield of 9.1% when progressing from nondiagnostic ES to GS^13^.

In this study, we applied singleton GS with expanded variant analyses to a large, comprehensively phenotyped DEE cohort unsolved after genetic testing. We aimed to assess the extent to which GS can close the diagnostic gap by expanding analysis beyond coding regions to include structural variants, intronic splice-altering variants, and non-coding gene variants.

## MATERIALS AND METHODS

### Cohort

The cohort included individuals with DEEs of unknown cause who satisfied the following criteria: (i) no obvious acquired cause, (ii) negative on any prior genetic testing, and (iii) DNA sample available for sequencing or genome data for reanalysis. Clinical data for each patient included a detailed medical history including seizure types, developmental course, all morbidities, examination findings, past medical, birth and family history. Additional information analysed included medical records, neuroimaging and EEG reports, and a validated seizure questionnaire^14^. Individuals were classified into specific electro-clinical syndromes according to the International League Against Epilepsy criteria where possible.

This project was approved by the Austin Health Human Research Ethics Committee (Project H2007/02961) and the WEHI Human Research Ethics Committee (Project G20/01). Written informed consent was obtained from parents or legal guardians.

### Genome sequencing

DNA was extracted primarily from blood, with saliva used where blood was unavailable. DNA was submitted for PCR-free GS at either Novogene (Beijing, China) or the Murdoch Children’s Research Institute (Melbourne, Australia) using the Illumina NovaSeq X Plus, 6000 or NextSeq 500 (Illumina, San Diego, United States) sequencers targeting an average read depth of 30x.

### Variant calling

Sequencing reads were aligned to the GRCh38 reference genome using BWA-MEM (v0.7.17). Short variants (SNVs and indels) were called following GATK short variant discovery best-practices guidelines using GATK4^15^ (v4.1.9), comprising base quality score recalibration (GATK BaseRecalibrator), germline short variant calling (GATK HaplotypeCaller), joint variant calling (GATK GenotypeGVCFs), and variant quality score recalibration (GATK VariantRecalibrator). Structural variants were called with multiple tools: the read-depth based caller CNVnator^16^ (v0.4.1), and the split-read/read-pair based callers Manta^17^ (v1.6.0) and smoove (v0.2.5).

### Variant annotation and prioritisation

Variants were annotated with gene features and consequences using Ensembl VEP^18^ (v110), with structural variant allele frequencies added with the StructuralVariantOverlap plugin based on the gnomAD structural variant database (v2.1). Short coding variants and structural variants were filtered and prioritised using an in-house pipeline (nf-cavalier, https://github.com/bahlolab/nf-cavalier), which implements filtering based on gene panels, predicted variant consequences, and population allele frequencies (<0.001 in gnomAD v4.1). The Genes4Epilepsy^6^ (v24-03) gene panel was used to restrict analyses to known monogenic epilepsy genes. Variants classified as pathogenic or likely pathogenic in ClinVar were considered regardless of whether the affected genes were present on the Genes4Epilepsy panel. Candidate structural variant calls were further filtered to reduce false positives by: 1) only retaining calls supported by multiple callers, and 2) manual review with the visualisation tool SVPV^19^. Putative splicing variants were further filtered using a combination of tools: SpliceAI^20^, Pangolin^21^, and CADD-Splice^22^, with any variant supported by multiple tools analysed further. Variants in non-coding small nuclear RNA (snRNA) genes recently implicated in epilepsy^23,24^ were shortlisted for further analysis if they were absent or rare in gnomAD v4.1.

### Variant classification and interpretation

Variants were classified according to the American College of Medical Genetics (ACMG) guidelines^25^. Pathogenic or likely pathogenic variants were validated and segregated in relevant family members where DNA was available. Variants of unknown significance were investigated further and reclassified as pathogenic or likely pathogenic based on additional evidence from: (i) segregation where possible to confirm *de novo* status in the proband, (ii) functional assays to confirm loss- or gain-of-function, and (iii) phasing of biallelic variants to show they were inherited *in trans*. Candidate variants were assessed against ClinVar classifications to determine whether they represented recurrent pathogenic alleles or were located within known mutational hotspots.

### Variant validation

#### Sanger Sequencing

Familial segregation of variants was established where possible. SNVs and indels were validated by PCR and Sanger sequencing. Amplification reactions were cycled using a standard protocol on a Veriti Thermal Cycler (Applied Biosystems, Carlsbad, CA). Bidirectional sequencing was completed with a BigDyeTM v3.1 Terminator Cycle Sequencing Kit (Applied Biosystems), according to the manufacturer’s instructions. Sequencing products were resolved using a 3730xl DNA Analyzer (Applied Biosystems).

#### Droplet digital PCR

Copy number changes associated with structural variants were validated by droplet digital PCR (ddPCR) with gene specific custom primers (Bioneer Pacific; sequences available on request). Reactions were prepared using QX200™ ddPCR™ EvaGreen Supermix (Bio-Rad) at a primer concentration of 0.1 µM with 10 ng DNA, in a final reaction volume of 22 µL. Droplets were generated using the manual QX200 Droplet Generator (Bio-Rad). PCR amplification was performed on a C1000 Touch thermal cycler (Bio-Rad) following the manufacturer’s recommendations. Droplet fluorescence was measured on the QX200 droplet reader, and data analysis was performed using QuantaSoft software (Bio-Rad).

#### Complementary DNA conversion for splicing assay

Blood samples were collected in PAXgene Blood RNA Tubes (Qiagen, Venlo, Netherlands), and RNA isolation was completed using the QIAamp RNA Blood Mini Kit (Qiagen) as per the manufacturer’s instructions. Isolated RNA was transcribed into complementary DNA (cDNA) using the SuperScript III Reverse Transcriptase kit (Invitrogen, Waltham, United States). Briefly, the first strand of cDNA was generated using a reaction mix consisting of 500 ng RNA, 50 ng random hexamers, 1 μL annealing buffer and ultrapure water up to a total of 8 μL on a Veriti 96 well Thermocycler (Applied Biosystems, California, USA) for 5 minutes at 65°C. Following this, 10μL of 2x First-Strand Reaction mix and 2μL of SuperScript III/RNAseOUT Enzyme mix was added and reactions cycled for 10 minutes at 25°C, 50 minutes at 50°C, and 5 minutes at 85°C.

#### Long-read sequencing

Long-read sequencing of blood-derived DNA was completed using the Ligation Sequencing DNA V14 kit (SQK-LSK114; Oxford Nanopore Technologies, UK) with R10.4.1 flowcells on a PromethION 2 Integrated platform, following the manufacturer’s protocols. Raw signal data were basecalled on-device using Dorado (server v7.6.7) with the super-accurate basecalling model (v4.3.0, 400 bps, 5hmC & 5mC CpG modifications enabled). The resulting reads were mapped to the GRCh38 human reference genome using minimap2 (v2.28-r1209). The EPI2ME Human Variation pipeline (v2.7.3) was applied to the aligned data, running Clair3 (v1.0.8) for small variant calling and WhatsHap (v2.5) for variant phasing and haplotype partitioning, enabling determination of whether co-occurring variants resided on the same chromosome (cis) or opposing chromosomes (trans).

### DNA methylation data analysis

Methylation analysis was performed to support a putatively pathogenic *MORC2* finding using the clinically validated EpiSign^TM^ laboratory workflow, following previously established methods^26^. Methylated and unmethylated signal intensities generated from the EPIC V1 arrays were imported into R 4.2.1 for normalization, background correction, and probe-level filtering. Beta values were calculated as a quantitative measure of methylation level, ranging from 0 (no methylation) to 1 (complete methylation).

Processed beta values from the case sample were compared with the established episignature for *MORC2* using the corresponding set of disorder-specific differentially methylated probes. Comparative analyses included hierarchical clustering and multidimensional scaling to assess concordance between the case sample and the reference cohort. These analyses enabled visualization of the case sample’s position relative to affected and control profiles within the established episignature framework. Final interpretation was based on the concordance of these comparative analyses.

## RESULTS

### Clinical Cohort

Genome sequencing (GS) was performed for 242 individuals (108 female) with unsolved DEEs. The median age at seizure onset for patients was 20 months (1 day – 18 years), and the median age at time of analysis was 24 years (range 6 months – 72 years). The cohort included 19 deceased individuals. The most common DEE syndromes were: 1) LGS in 56 patients (23%); 2) EMAtS in 45 cases (19%); 3) DEE-SWAS in 21 cases (9%); and 4) IESS in 20 cases (8%).

Most participants (92%, 223/242) had undergone prior multigene sequence-based testing: 30% (71/242) had multigene panel (MGP) testing only – primarily via molecular inversion probe assays^27^, 16% (39/242) had exome sequencing (ES) only, and 47% (113/242) had both. Most participants had also undergone prior chromosomal microarray (CMA) testing 74% (178/242).

### Diagnostic Yield

Pathogenic or likely pathogenic (P/LP) variants were identified for 36 participants (15%), and variants of uncertain significance for 10 participants (4%) (Figure 1, Table 1, Tables S1). The majority of P/LP variants identified were short variants (SNVs/Indels) in coding regions (n=26), followed by structural variants (deletions/duplications, n=9), and lastly short variants in non-coding regions (n=5). Three participants had compound heterozygous P/LP variants, and one participant (P7) had P/LP variants in two genes (*CACNA1A* and *PCDH19*).

**Figure 1:**
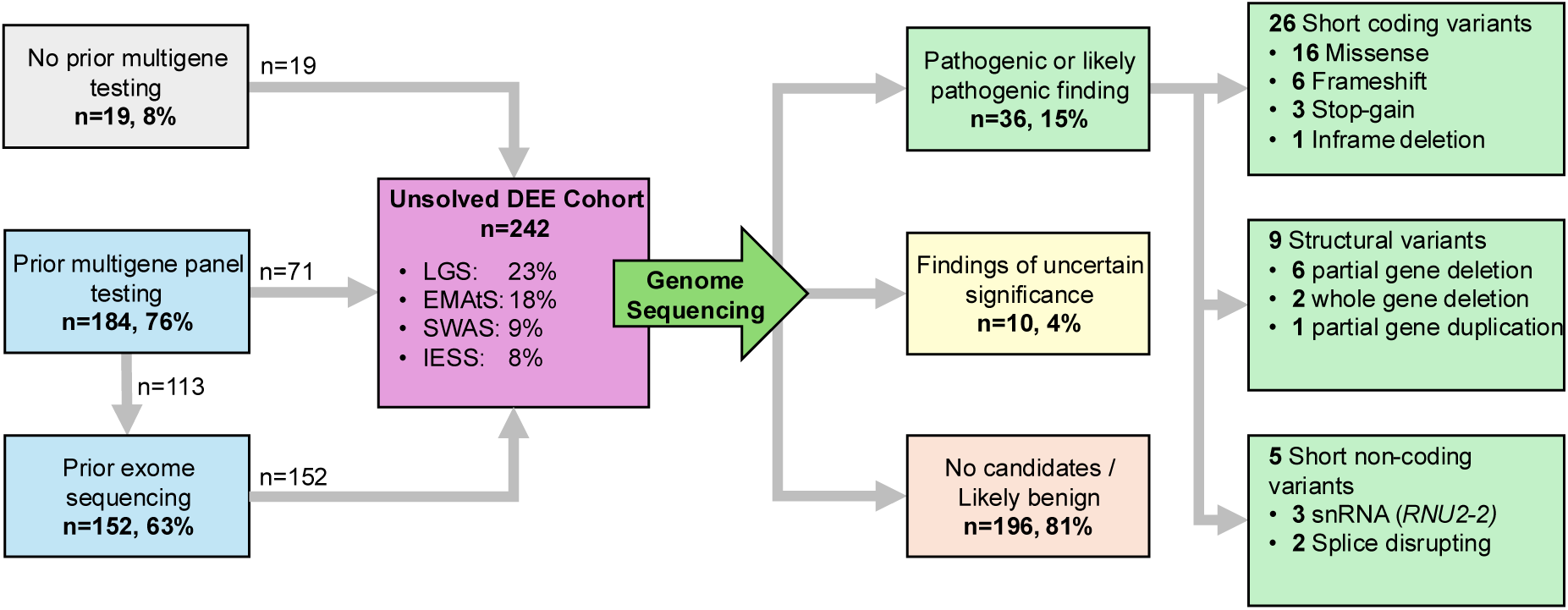
Schematic overview of unsolved DEE genome sequencing study. Note that the number of participants with P/LP findings (n=36) is less than the total number of P/LP variants (n=40) as three participants had compound heterozygous variants (i.e. two distinct variants per participant) and one had P/LP in two different genes. LGS = Lennox-Gastaut syndrome, EMAtS = epilepsy with myoclonic-atonic seizures, SWAS = developmental and epileptic encephalopathy with spike-wave activation in sleep, IESS = infantile epileptic spasms syndrome, snRNA = small nuclear RNA.

**Table 1:**
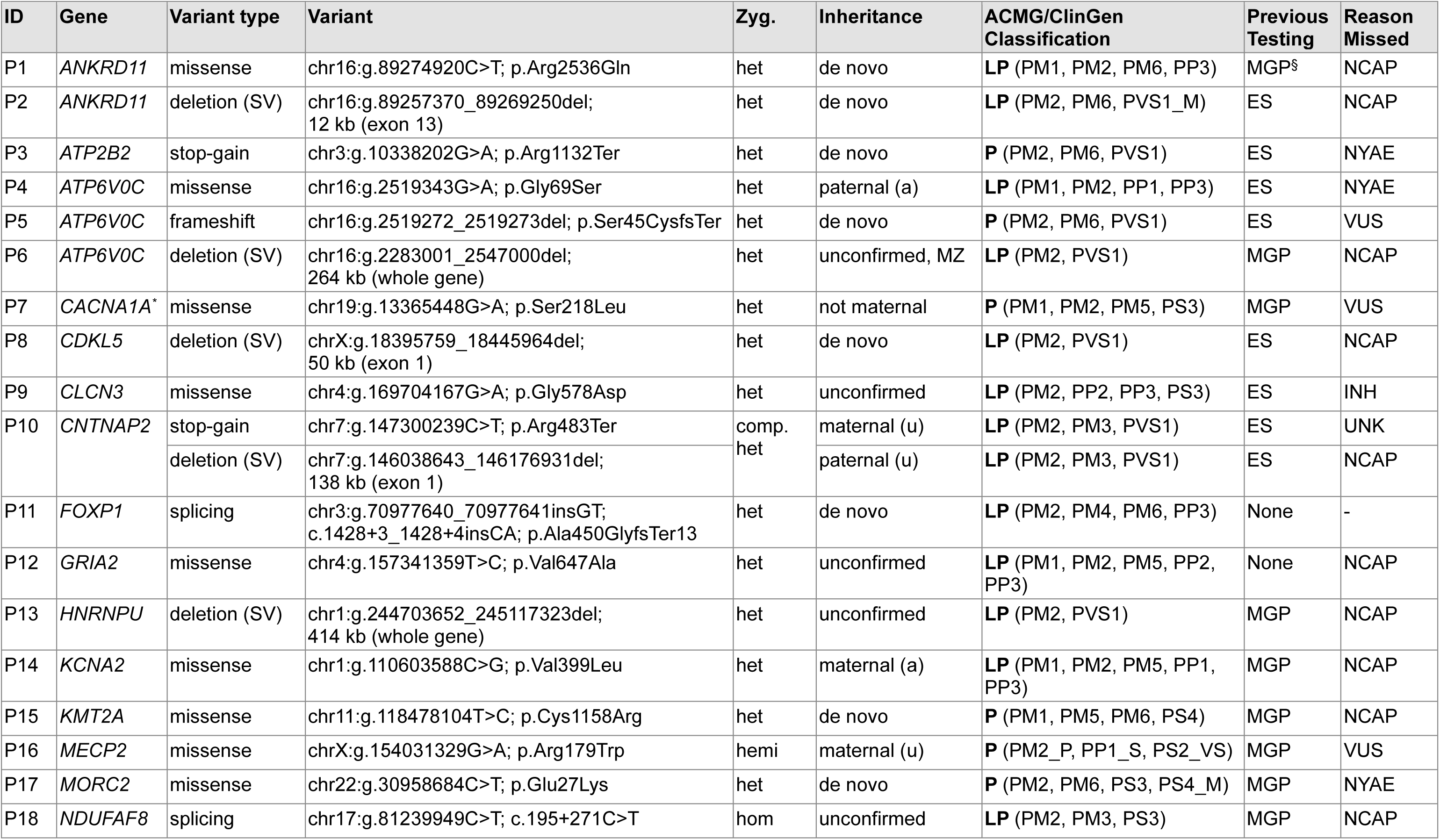
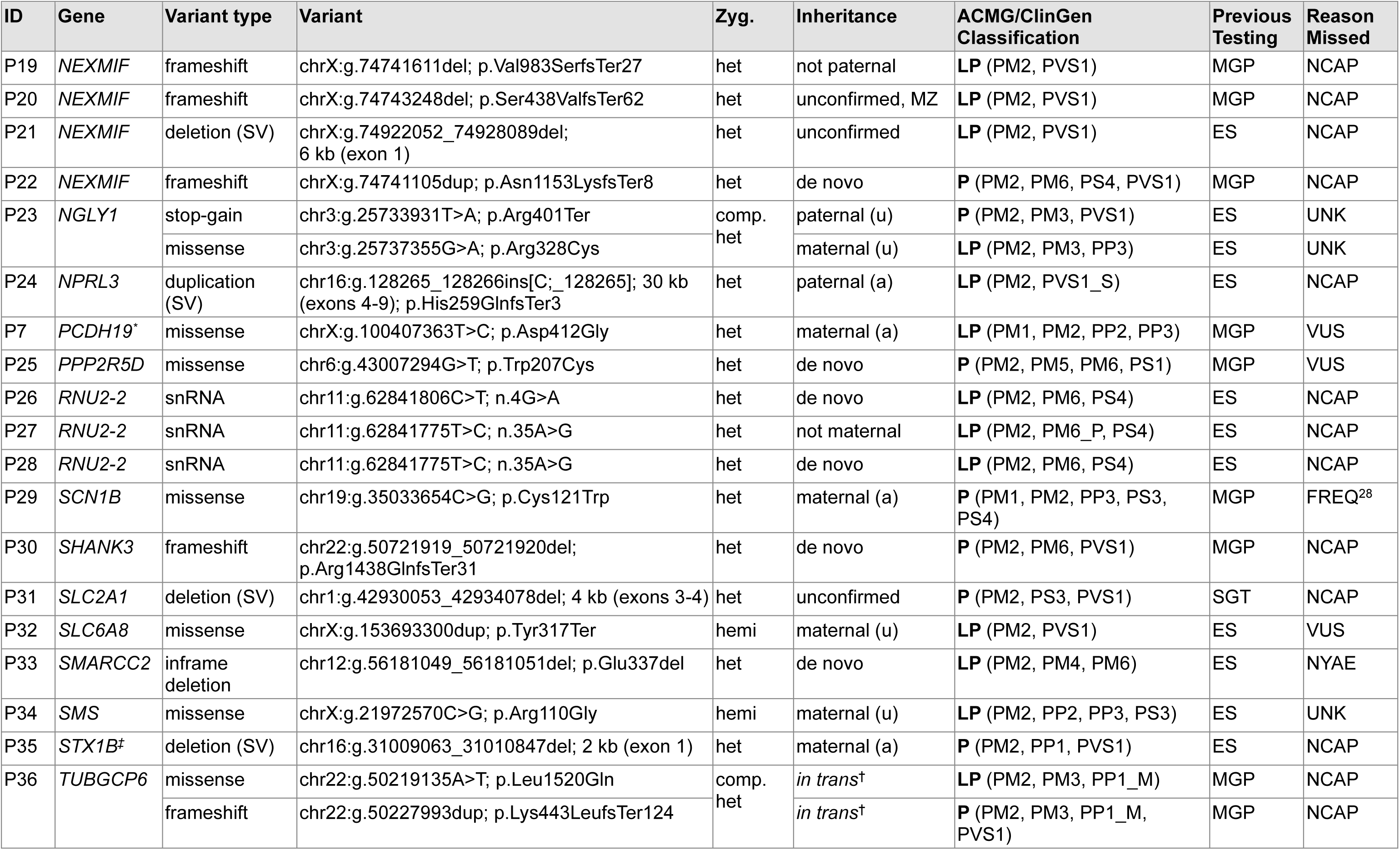
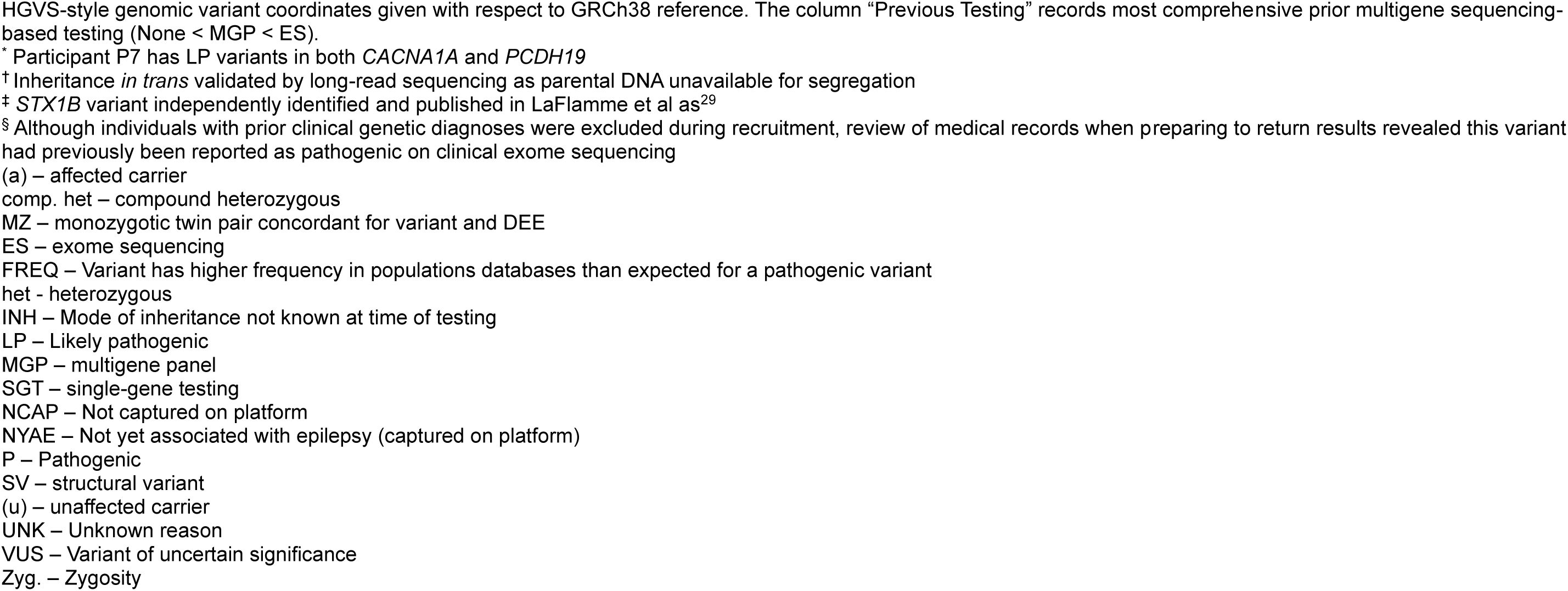
Pathogenic and likely pathogenic variants identified in our cohort.

Diagnostic yields were examined based on history of prior testing. Cases with no prior MGP or ES had a yield of 10% (2/19, 95% CI 1-33%), lower than the expected 50% solve rate for test-naïve cases. However, this group was not strictly test-naïve: 37% (7/19) had undergone prior CMA testing and 16% (3/19) had family members with a similar presentation who had undergone sequence-based testing. Cases with prior MGP and without ES had a yield of 23% (16/71, 95% CI 13% - 34%), and those with prior ES had a yield of 12% (18/152, 95% CI 7% - 18%). A one-sided Fisher’s exact test indicated that participants who had undergone prior MGP testing only had a significantly higher rate of a molecular diagnosis on subsequent GS compared to those who had undergone prior ES (odds ratio=2.2, p=0.03), consistent with the more limited scope of MGP testing compared to ES.

Diagnostic yields were examined for patients with the most common DEE syndromes in our cohort: IESS 5% (1/20, 95% CI 0% - 25%), DEE-SWAS 5% (1/21, 95% CI 0% - 24%), LGS 14% (8/56, 95% CI 6% - 26%), EMAtS 16% (7/45, 95% CI 6% - 29%), and all other cases had a yield of 19% (22/114, 95% CI 13% - 28%). There were no significant differences in diagnostic yield based on pairwise two-sided Fisher’s exact tests comparing DEE syndromes.

### Structural variants

P/LP structural variants (excluding indels less than 50 bp) were identified in 9/242 (3.7%) patients and accounted for 9/40 (23%) P/LP variants (Table 1). These comprised eight deletions and one duplication, all of which were undetectable by MGP or ES. Their sizes ranged from 2 kb (*STX1B* exon 1 deletion) to 414 kb (*HNRNPU* whole gene deletion).

Participant P31, who had biochemical findings consistent with GLUT1 deficiency but was negative for targeted *SLC2A1* sequencing, harboured a 4 kb deletion encompassing exons 3–4 of *SLC2A1*, including the functionally crucial transmembrane domain TM4^30^ (Table 1, Figure 2A). Participant P13 had a 414 kb deletion spanning *HNRNPU*. She had features consistent with *HNRNPU*-related neurodevelopmental disorder, including early-onset seizures, tonic-clonic and absence seizures, mild-moderate intellectual disability, short stature, hypotonia and scoliosis^31^. Participant P35 had a heterozygous 1.8 kb exon 1 deletion of *STX1B*, and was identified orthogonally with a differentially methylated region upstream of *STX1B* in LaFlamme et al^29^. Participant P10 had a heterozygous 138 kb deletion removing the first exon of *CNTNAP2*, which is associated with the autosomal recessive Pitt-Hopkins-like syndrome-1. The participant also carried a stop-gain variant in *CNTNAP2,* and segregation confirmed compound heterozygous inheritance. Participant P2 had a 12kb deletion of exon 13 of *ANKRD11,* which was independently identified from a clinical high-density CMA. Segregation revealed *de novo* inheritance. He had features consistent with KBG syndrome with seizures during childhood, mild ID, short stature, macrodontia and missing teeth, right strabismus, dysmorphic facial features (synophrys, widely spaced eyes, broad eyebrows, prominent nasal bridge, bulbous nose, anteverted nares), fifth digit brachydactyly and foetal finger pads.

**Figure 2:**
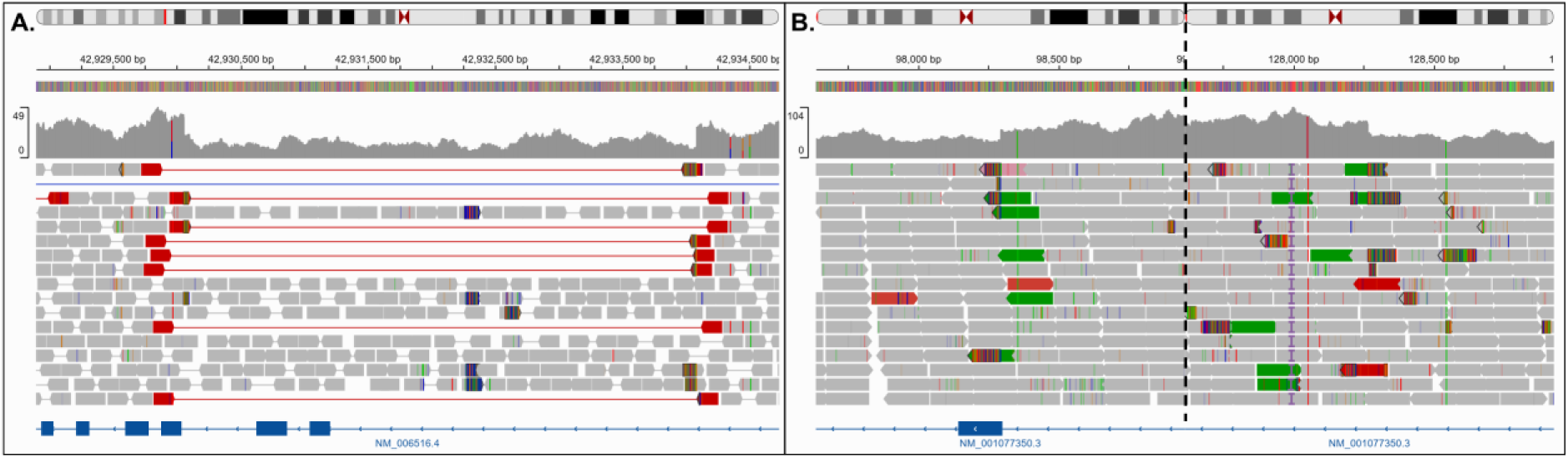
IGV snapshots of selected structural variants identified in our cohort. A) Heterozygous deletion of *SLC2A1* exons 3-4 in participant P31. Reads connected by red lines indicated read-pairs that span the deletion. B) Heterozygous tandem duplication within *NPRL3* in participant P24, causing incomplete duplication of exons 4-8 and partial duplication of exon 9 resulting in a predicted frameshift. The dashed black line represents discontinuity between the left and right breakpoint visualisations; green reads indicate the inverted read-pair orientation characteristic of tandem duplication breakpoints.

Participant P24 carried a heterozygous duplication spanning exons 4-9 of *NPRL3,* with a breakpoint in exon 9 predicted to cause a frameshift (Figure 2B). *NPRL3* is associated with focal epilepsy^32^, and our patient had a DEE with focal seizures in early childhood. Developmental plateauing was associated with seizure onset and she now presents with mild-moderate ID. Her MRI identified multiple cortical malformations including cortical dysplasia, polymicrogyria, and periventricular heterotopia, consistent with *NPRL3*-associated epilepsy, as well as subcortical band heterotopia, which is not typically associated with *NPRL3*. The variant was inherited from the participant’s unaffected father, however incomplete penetrance is seen in *NPRL3*^33^.

### Non-coding variants

P/LP non-coding variants were identified in 5/242 (2%) participants and accounted for 5/40 (13%) P/LP variants (Table 1). These included two splice-disrupting variants and three variants in the small nuclear RNA (snRNA) gene *RNU2-2*. Of the five, only one splice-disrupting variant could have been detectable by ES, as it lay within ten base pairs of an exon boundary, whereas the other splice-disrupting variant was deep intronic, and snRNA genes are not captured by ES platforms.

The first splice-disrupting variant was identified in participant P11 in *FOXP1*. The c.1428+3_1428+4insCA variant was predicted by SpliceAI to cause exon 16 to be skipped (donor loss Δ score = 0.98 at +3, acceptor loss Δ score = 0.88 at +79), resulting in a frameshift. This variant arose *de novo* and exon-16 skipping was confirmed in blood-derived RNA by ddPCR using primers targeting the mis-spliced exon-15/exon-17 junction, and the canonical exon-17/exon-18 junction.

A second splice-disrupting variant was identified through a genome-wide screen for recurrent pathogenic variants reported in ClinVar. While most variants were interrogated because they were in genes on the Genes4Epilepsy panel, this analysis highlighted *NDUFAF8*, which was not on the panel at the time of analysis. Participant P18 was homozygous for the deep intronic *NDUFAF8* c.195+271C>T variant, as was his similarly affected sister. Biallelic loss-of-function variants in *NDUFAF8* have been associated with Leigh syndrome, a mitochondrial disorder that includes seizures, and the c.195+271C>T variant abolishes expression of the functional *NDUFAF8* isoform^34^. Our siblings and the 3 reported patients with this variant have the unifying features of IESS, ID, visual problems and elevated blood lactate.

We identified three individuals with recurrent *de novo* pathogenic variants in the snRNA gene, *RNU2-2*, one with the n.4G>A variant and two with the n.35A>G variant. These *RNU2-2* variants cause neurodevelopmental disorders^23,35^. Their detailed phenotypic data contributed to our recent delineation of the features of *RNU2-2* DEE: seizure onset at a median age of 24 months, multiple seizure types, high rates of status epilepticus, drug resistance, and notable features of hyperventilation and obstructive sleep apnoea^36^.

### Short coding variants

P/LP short variants in coding regions were identified in 23/242 (9.5%) participants and accounted for 26/40 (65%) P/LP variants (Table 1). All of these variants are theoretically detectable on ES, however, many of the affected genes were not robustly associated with epilepsy at the time of previous analysis. Three genes were identified across multiple patients: four had variants in *NEXMIF* (3 frameshifts, one structural variant), three had variants in *ATP6V0C* (including one structural variant), and two had variants in *ANKRD11* (including one structural variant). The remaining genes were only found in one patient.

Participant P17 harboured a *de novo MORC2* p.Glu27Lys variant identified through the ClinVar recurrent variant screen. Pathogenic variants in *MORC2* are associated with a neurodevelopmental disorder that includes axonal neuropathy and features suggestive of mitochondrial disease^37^. The clinical features in participant P17 overlap with those reported in the five previously described individuals carrying the same p.Glu27Lys variant, including short stature (5/5), microcephaly (3/5), hearing loss (2/5), hypotonia (2/5), and epilepsy (1/5)^37^. *MORC2* has an established role in epigenetic silencing^38^, and we ascertained that participant P17 was concordant for a *MORC2*-associated genome-wide methylation profile (episignature) (Figure 3).

**Figure 3:**
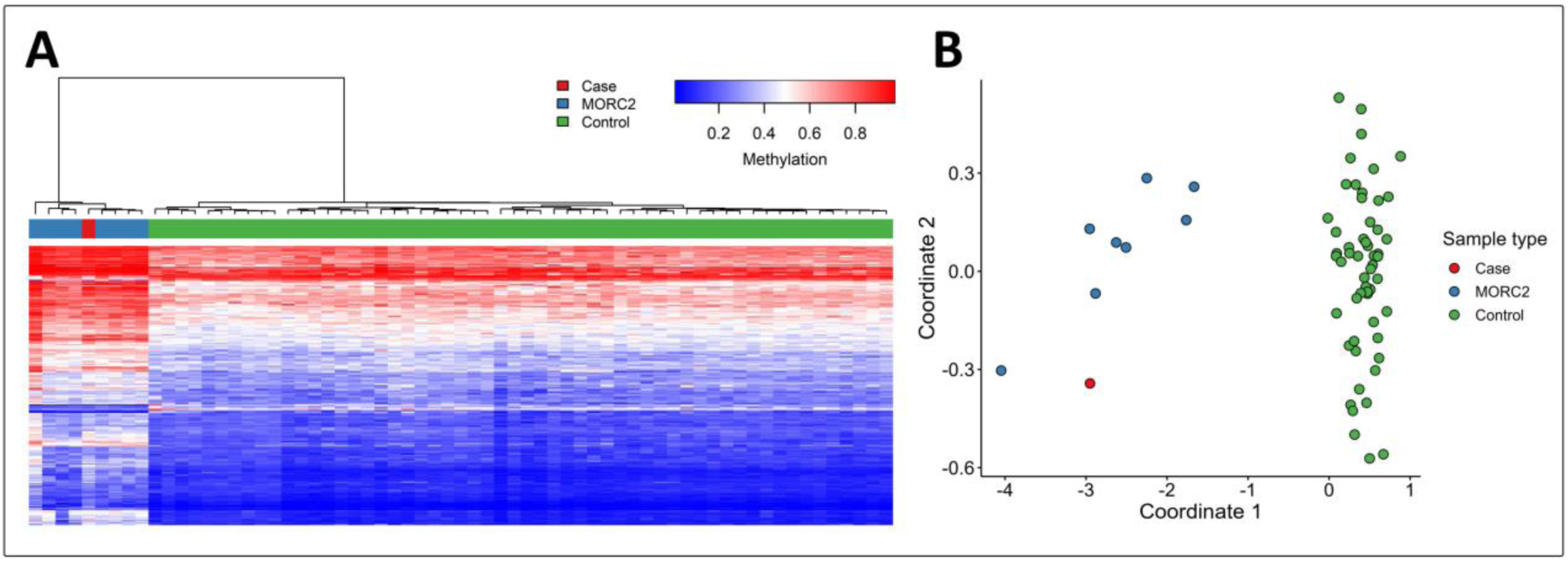
DNA methylation analysis for the *MORC2* episignature. A. Heatmap with hierarchical clustering of disorder-specific differentially methylated probes shows that the case sample (red) clusters with individuals carrying the established *MORC2* episignature (blue) and separate from unaffected controls (green). Each row represents a differentially methylated probe, and each column represents a sample. B. Multidimensional scaling analysis demonstrates the same pattern, with the case positioned within the *MORC2* cluster and distinct from controls, consistent with concordance to the *MORC2* methylation episignature.

A *de novo* stop gain in *ATP2B2* (p.Arg1132Ter) was determined to be likely pathogenic for participant P3. The gene was not present on the analysis version of Genes4Epilepsy (v24-03) and was identified through a novel machine learning model “PanRank” trained on the Genes4Epilepsy list (Munro et al., in preparation, bahlolab.github.io/PanRank). The gene was also present on the PanelApp Australia “Genetic Epilepsy” panel and has since been included in Genes4Epilepsy. *De novo* loss-of-function *ATP2B2* variants are associated with neurodevelopmental disorders featuring epilepsy, dystonia, ataxia, and intellectual disability^39^.

## DISCUSSION

Applying GS to an unsolved cohort of individuals with DEEs yielded a diagnostic rate of 15%, increasing the yield substantially from ES or MGP. Given that 92% of our patients had undergone prior multigene testing, this cohort is enriched for cases difficult to solve on standard genetic testing. The yield of genetic testing will grow as our understanding and ability to interpret structural variants and non-coding variants improves, driven by novel bioinformatic approaches, annotation resources and sequencing platforms which capture previously missed variation.

Structural variants and non-coding variants accounted for almost one-third of the diagnostic variants identified in our cohort. These variants are broadly inaccessible to MGP and ES and underscore the key role of GS in increasing the rate of molecular diagnoses for the DEEs. All structural variants identified were copy-number variants (CNVs) smaller than 500 kb, below the detection threshold of standard CMA. Although high-density CMA platforms can now detect smaller events, as illustrated by the 12 kb *ANKRD11* deletion identified independently by both GS and an external clinical high-density CMA for participant P2, such platforms are not yet widely implemented in clinical practice. Furthermore, without accurate breakpoint resolution, it is impossible to accurately predict the consequences of CNVs with intragenic breakpoints using CMA as the precise gene disruption is not apparent.

The remaining two-thirds of genetic diagnoses were largely driven by the expanded knowledge of causative genes for DEEs. Over the past decade, large-scale sequencing efforts have greatly expanded the catalogue of genes associated with epilepsy and neurodevelopmental disorders. Consequently, variants in some genes (i.e., *ATP2B2*, *MORC2*) that were uninterpretable at the time of prior testing, have now been recognized as pathogenic. Curated and regularly updated open-access gene panels, such as Genes4Epilepsy and PanelApp, have become invaluable tools for the detection, interpretation and (re-)classification of disease-associated variants.

Routine reanalysis of ES/GS data yielded additional molecular diagnoses in suspected Mendelian diseases, with a meta-analysis of 29 studies across many diseases reporting an overall diagnostic gain of approximately 10% and recommending reanalysis after two years unless clinically indicated^40^. In principle, reanalysis of GS offers greater potential than ES as it captures structural and non-coding variants, providing a more comprehensive set of variants for reinterpretation. Advances in variant effect prediction, particularly those employing deep learning to predict the effect of non-coding variants on splicing and gene regulation (e.g. SpliceAI^20^), are beginning to enable the systematic prioritisation and evaluation of non-coding variants captured by GS for confirmation at the RNA level. As these tools mature and become embedded in variant interpretation frameworks, reanalysis of GS data will increasingly yield new molecular diagnoses.

Despite its many advantages, short-read GS still has some limitations. Certain variant classes, such as long repeat expansions, mobile-element insertions, complex structural rearrangements, and variants in low-complexity or highly repetitive genomic regions, may be incompletely resolved. Long-read genome sequencing (LRS) can overcome many of these limitations. A recent study reported an increased diagnostic yield of approximately 7% when applying long-read GS to a neurodevelopmental disorder (NDD) cohort negative on short-read GS^41^. As the sequence error rate of clinical LRS platforms reduces and costs decrease, LRS will increasingly complement and may eventually supersede short-read GS in diagnostic workflows. Additionally, the availability of methylation data through LRS is likely to increase the diagnostic rate as more episignatures for NDDs are identified, though progress for DEEs specifically has been limited, with positive episignatures largely found in NDDs associated with DEEs^29^.

This study has demonstrated the utility of GS in providing molecular diagnoses for a large cohort of patients with DEEs unsolved on standard genetic testing. Detection of structural variants and non-coding variants undetectable on ES or MGP accounted for a substantial proportion of the new diagnoses. Broader implementation of GS as a first-line test will help close the diagnostic gap, guiding management, prognostic and genetic counselling, and provide a solid base enabling future reanalysis for cases that remain unsolved.

## Data Availability

The raw data that support the findings of this study are not publicly available due to ethical restrictions and participant consent limitations. The dataset contains sensitive genomic information from individuals with rare disease, for which informed consent was obtained only for use within the scope of this study. Requests for access to de-identified data will be considered on a case-by-case basis, subject to institutional ethics approval and a formal data sharing agreement. Enquiries should be directed to the corresponding author.

## SUPPLEMENTARY INFORMATION

**Supplemental Table 1:**
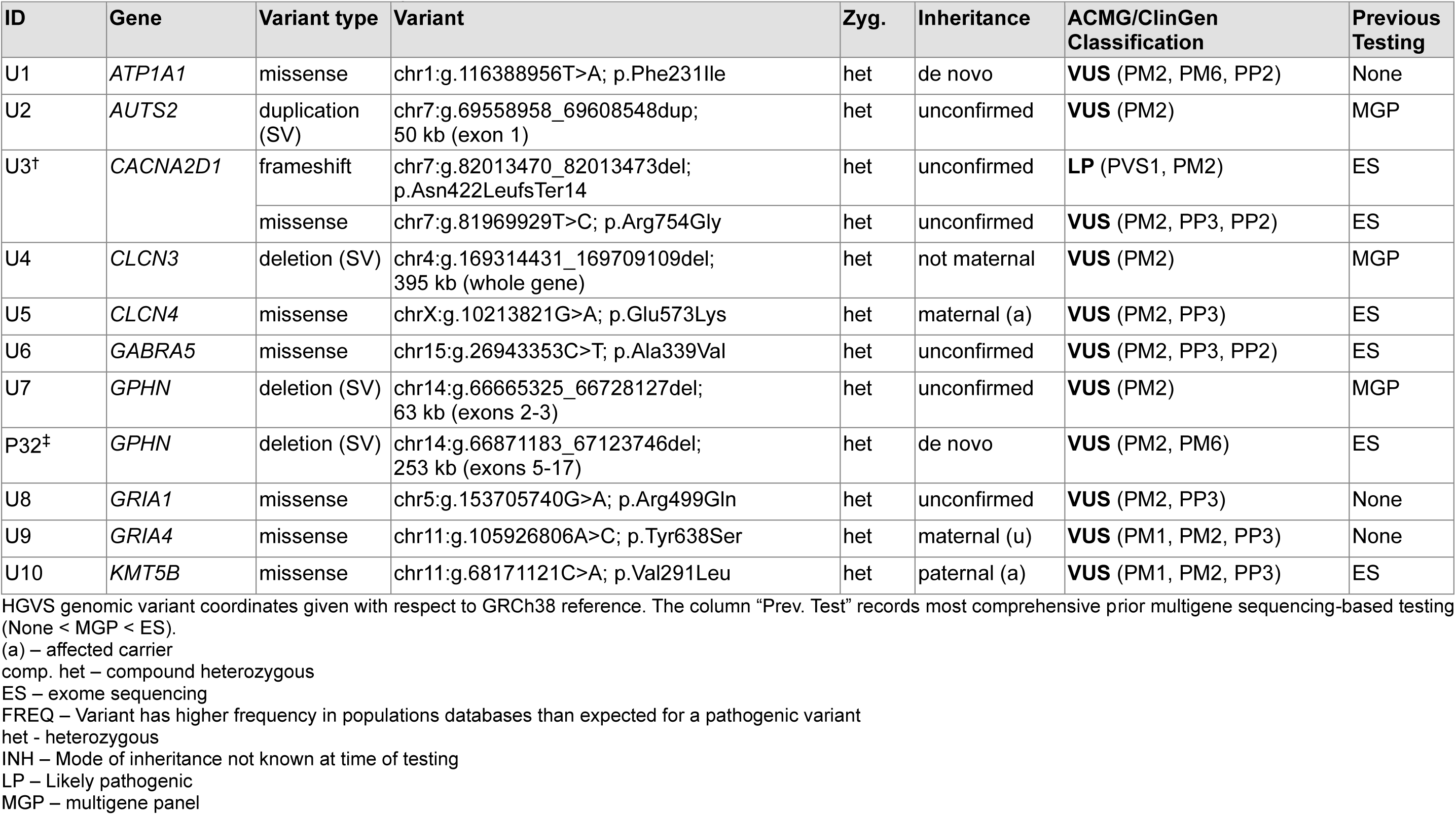
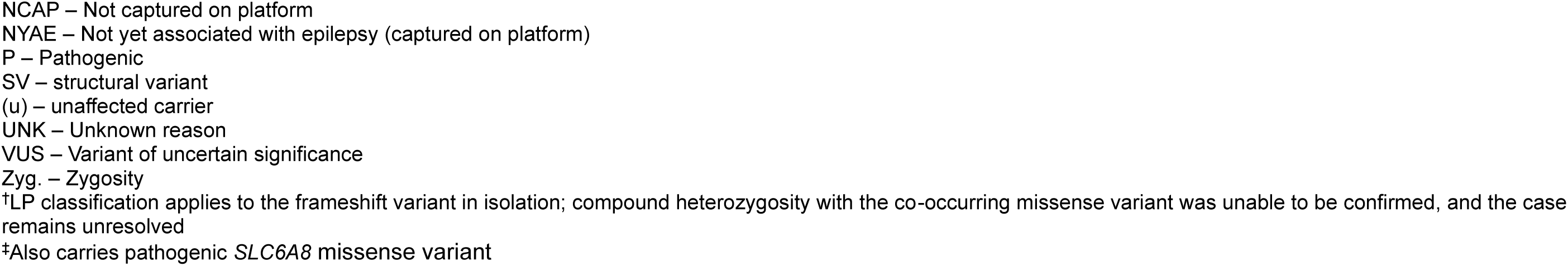
Variants of uncertain significance identified in our cohort.

